# Neural Closed Loop Deep Brain Stimulation for Freezing of Gait

**DOI:** 10.1101/19011858

**Authors:** Matthew N. Petrucci, Raumin S. Neuville, M. Furqan Afzal, Anca Velisar, Chioma M. Anidi, Ross W. Anderson, Jordan E. Parker, Johanna J. O’Day, Kevin B. Wilkins, Helen M. Bronte-Stewart

## Abstract

Freezing of gait (FOG), a devastating symptom of Parkinson’s disease (PD), can be refractory to current treatments such as medication and open-loop deep brain stimulation (olDBS). Recent evidence suggests that closed-loop DBS (clDBS), using beta local field potential power from the subthalamic nucleus (STN) as the control variable, can improve tremor and bradykinesia; however, no study has investigated the use of clDBS for the treatment of FOG. In this study, we provide preliminary evidence that clDBS was superior to olDBS in reducing percent time freezing and in reducing freezing behavior (gait arrhythmicity) in a person with PD and FOG. These findings warrant further investigation into the use of clDBS to treat FOG while also minimizing the total energy delivered to maintain a therapeutic effect.

---

Dear Editor,

Freezing of gait (FOG) is a devastating symptom of Parkinson’s disease (PD), affecting over half of the patient population [1] and negatively impacting mobility and patient quality of life. This symptom has been difficult to treat with dopaminergic medication, is associated with arrhythmic gait, and can become refractory over time [2]. Moreover, it is debated to what extent deep brain stimulation (DBS) provided in an open-loop manner (olDBS) can mitigate FOG [3].

Neural closed-loop deep brain stimulation (clDBS) has been demonstrated to alleviate the signs and symptoms of PD by adjusting stimulation in response to elevations in local field potential (LFP) beta band (13-30 Hz) power in the subthalamic nucleus (STN). Improvements in tremor and bradykinesia on clDBS have been observed using beta power as the control variable in both single and dual threshold algorithms [4]–[6]. We have shown that STN olDBS attenuated pathological beta fluctuations while improving FOG [7]. To date, no study has used similar closed-loop paradigms to reduce FOG. In this paper, we demonstrate preliminary evidence that clDBS driven by STN beta band power was superior to conventional olDBS in reducing the percent time freezing and arrhythmicity during a stepping in place (SIP) task.

One male participant (age: 63.2 years, off UPDRS-III: 55, disease duration: 5.1 years, akinetic-rigid subtype) with PD and FOG participated in the study. The participant was implanted with an investigative sensing neurostimulator (Medtronic Activa^®^ PC+S, FDA IDE approved) and bilateral STN DBS leads (Medtronic model 3389). All procedures were approved by the Stanford University Institutional Review Board and the participant provided informed written consent.

The participant performed the SIP task [8] during four stimulation conditions: off DBS (OFF), on his clinical open-loop settings (c-olDBS), on voltage matched open-loop DBS (m-olDBS), and on closed-loop DBS (clDBS) (see Table S1 for the stimulation parameters). The stimulation settings for m-olDBS were the same single contacts used in the clDBS condition set to the average voltage calculated during clDBS. All testing was performed in the off-medication state (refrained for 12 hours for short- and 24/48 hours for long-acting dopaminergic medication). The clDBS was modulated by the power of local field potentials contained in the beta frequency range [4]. The maximum voltage that provided clinical improvement without side effects (V_Max_) in each STN was determined (left STN: 4.3 V, right STN: 4.5 V). The dual threshold control algorithm parameters were determined from beta band power during movement. This “movement band” beta power was measured during voltage titration SIP trials at 5 voltages between 0 and 100% of V_Max_ presented in random order (Figure S1). The “movement band” was set to ±3 Hz around the peak frequency of elevated beta band power during SIP (15 Hz for both STNs) [7]. The upper and lower values of the dual threshold controller were set to the average beta power measured during the stepping in place task at the minimum voltage (V_Min_) that showed improvement in stepping and freezing behavior (upper beta threshold) and at V_Max_ (lower beta threshold); V_Min_ was 25% of V_Max._ Previously established ramp rates were used for both STNs [9] (0.1 V/0.4 s up, and 0.1 V/0.8 s down).

An automated algorithm detected freezing events when the participant’s feet did not lift off the force plates [8]. FOG and freezing behavior were assessed using the percent time freezing and arrhythmicity (coefficient of variation (CV) of stride time), respectively. Total electrical energy delivered (TEED) and volume of tissue activated (VTA) was calculated for all stimulation conditions (see supplemental methods).

In the OFF-DBS condition the subject exhibited FOG at the start of the stepping task, was able to step for ~ 20 seconds and then experienced prolonged FOG as his repetitive stepping behavior deteriorated (i.e., loss of force modulation, Fig. 1A). While on c-olDBS and m-olDBS, there was improvement in the duration of normal stepping but FOG episodes were still detected (Fig. 1B and C). During clDBS, only a short start hesitation was detected at the beginning of the episode (Fig. 1D). The percent time freezing was 68.7% OFF DBS, 2.3% on c-olDBS, 23.5% during m-olDBS, and 1.5% during clDBS. SIP arrhythmicity was lower in all stimulation conditions compared to OFF (54.9% OFF, 18.2% c-olDBS, 27.4% m-olDBS, 5.2% clDBS, Figs. 1A-D). There was an increase in arrhythmicity after the first 25 seconds of the trial during both c-olDBS and m-olDBS, but during clDBS, stepping remained rhythmic (Fig. 1E). There was no difference in TEED between m-olDBS and clDBS, and the average TEED was 2% higher in clDBS vs. c-olDBS (Table S2).

**Figure 1:**
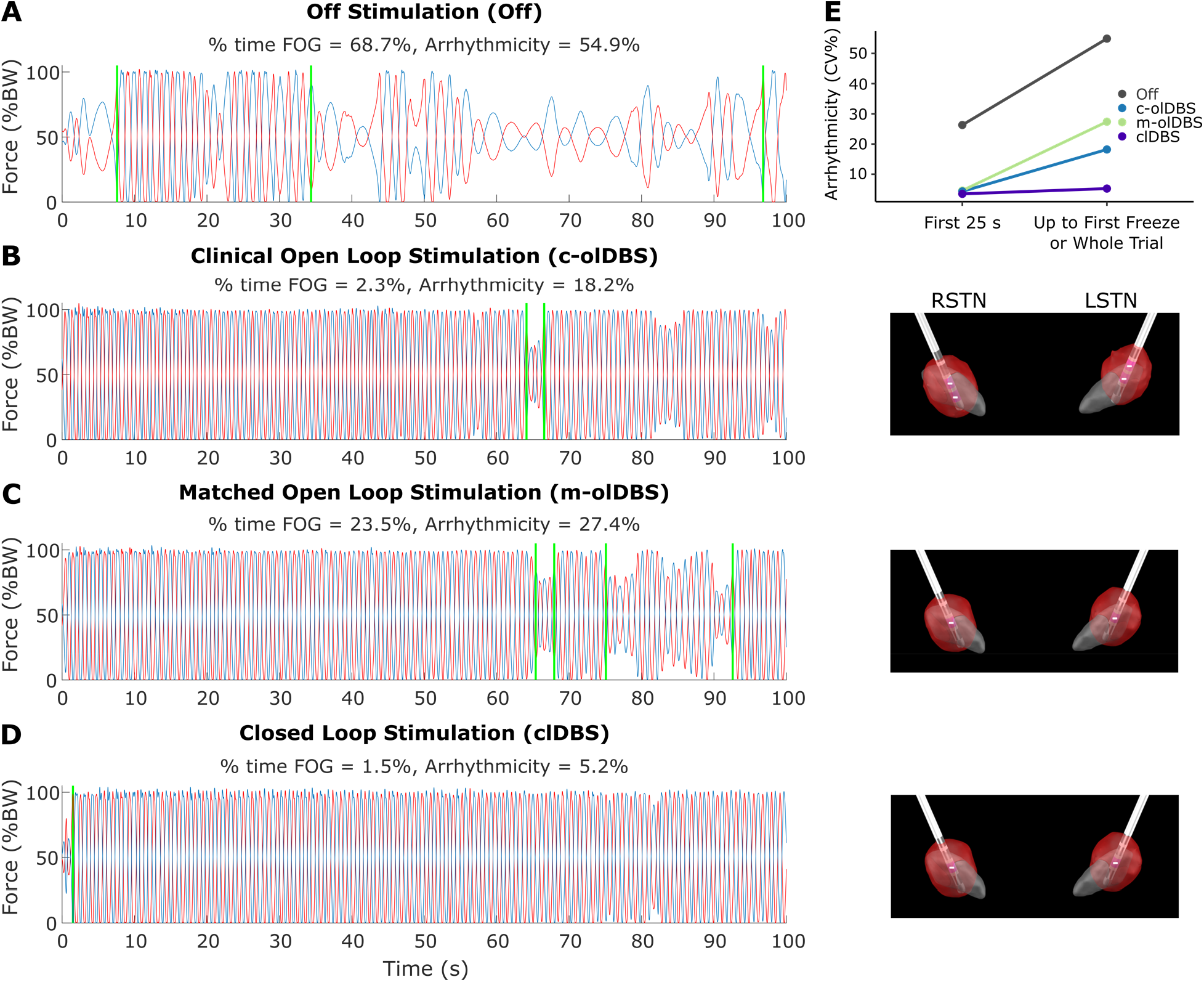
Stepping in place vertical ground reaction forces for the participant (**A**) off stimulation, (**B**) on clinical open loop stimulation, (**C**) on matched open loop stimulation, and (**D**) on neural closed loop stimulation. FOG events detected by automated algorithm [8] are indicated by the vertical green lines. Percent time freezing and arrhythmicity of the whole trial or up to the first freeze are presented above each condition. The volume of tissue activated from each STN is to the right of each condition in red. Although stimulation improved stepping in all conditions, closed loop stimulation showed the lowest arrhythmicity and % time freezing. Arrhythmicity (**E**) of the first 25 seconds and up to the first freeze or whole trial (if there were no freezing events) are plotted for each condition. Arrhythmicity was overall higher off stimulation and continued to worsen later in the trial for all conditions except for closed loop stimulation.

These findings, to the best of our knowledge, are the first to demonstrate that neural closed-loop DBS (clDBS), using a dual threshold algorithm based on beta power determined by therapeutic voltage titrations, was superior to clinical open loop DBS (c-olDBS), matched open loop DBS (m-olDBS), and no DBS (OFF) in reducing FOG in PD. Freezing behavior, manifesting as arrhythmic stepping and lack of maintaining a consistent rate of force/amplitude control during stepping (i.e., the “sequence effect” [10]), also improved more during clDBS compared to c-olDBS, m-olDBS, and OFF. Both c-olDBS and m-olDBS resulted in a similar deterioration of stepping behavior despite a small increase in TEED and VTA during m-olDBS. However, stepping behavior was maintained during clDBS even though the TEED and VTA were nearly identical to m-olDBS. These findings suggest that allowing the stimulation to adapt during the trial may allow the motor system to sustain or regain movement control, whereas continuous stimulation (with a similar or same amount of TEED and VTA) cannot prevent the “sequence effect” that contributes to arrhythmic gait and FOG [10] because it is not changing in response to fluctuating STN activity. Overall, these findings warrant further investigation into the use of clDBS for improving FOG as well as other Parkinsonian symptoms. Future investigations should evaluate how much the stimulation needs to adapt to maintain a therapeutic effect while also minimizing the energy requirement.

## Data Availability

N/A

## Acknowledgments

The authors would like to thank the participant in the study as well as members of the Human Motor Control and Neuromodulation Lab. This work was supported by the Michael J. Fox Foundation (9605), NINDS Grant 5 R21 NS096398-02, NIH Grant AA023165-01A1, Robert and Ruth Halperin Foundation, John A. Blume Foundation, Helen M. Cahill Award for Research in Parkinson’s Disease, the Stanford Bio-X Graduate Fellowship, and Medtronic Inc., who provided the device used in this study but no additional financial support.

## References

[1] M. Macht et al., “Predictors of freezing in Parkinson’s disease: a survey of 6,620 patients,” Mov. Disord., vol. 22, no. 7, pp. 953–956, 2007.

[2] N. Giladi, “Medical treatment of freezing of gait,” Mov. Disord., vol. 23, no. S2, pp. S482–8, 2008.

[3] M. U. Ferraye, B. Debu, and P. Pollak, “Deep brain stimulation effect on freezing of gait,” Mov. Disord., vol. 23 Suppl 2, pp. S489–94, 2008.

[4] A. Velisar et al., “Dual threshold neural closed loop deep brain stimulation in Parkinson disease patients,” Brain Stimul., vol. 12 no. 4, pp. 868–876, 2019.

[5] M. Arlotti et al., “Eight-hours adaptive deep brain stimulation in patients with Parkinson disease,” Neurology, vol. 0, p. 10.1212/WNL.0000000000005121, 2018.

[6] S. Little et al., “Adaptive deep brain stimulation in advanced Parkinson disease,” Ann. Neurol., vol. 74, no. 3, pp. 449–457, 2013.

[7] C. Anidi et al., “Neuromodulation targets pathological not physiological beta bursts during gait in Parkinson’s disease,” Neurobiol. Dis., vol. 120, no. May, pp. 107–117, 2018.

[8] J. Nantel, C. de Solages, and H. Bronte-Stewart, “Repetitive stepping in place identifies and measures freezing episodes in subjects with Parkinson’s disease,” Gait Posture, vol. 34, no. 3, pp. 329–333, 2011.

[9] M. F. Afzal, A. Velisar, C. Anidi, R. Neuville, V. Prabhakar, and H. Bronte-Stewart, “Proceedings #61: Subthalamic Neural Closed-loop Deep Brain Stimulation for Bradykinesia in Parkinson’s Disease,” Brain Stimul., vol. 12, no. 4, pp. e152–e154, 2019.

[10] R. Chee, A. Murphy, M. Danoudis, N. Georgiou-Karistianis, and R. Iansek, “Gait freezing in Parkinson’s disease and the stride length sequence effect interaction,” Brain, vol. 132, no. 8, pp. 2151–2160, 2009.

